# The effect of face mask mandates during the COVID-19 pandemic on the rate of mask use in the United States

**DOI:** 10.1101/2020.10.03.20206326

**Authors:** Michael J. Maloney

## Abstract

As COVID-19 continues to spread throughout the United States, there has been a search for policies to prevent individual infections, to slow the spread of the virus in general, and to mitigate the economic impact of the pandemic. Masks have proven to be a cost-effective measure in all regards, and as such some state governments have begun to mandate their use. However, while the efficacy of masks has been demonstrated, the efficacy of public policies which mandate the use of masks has not been demonstrated. This paper compares the rates of mask use in counties as defined by state policy. It finds that state mandates are strongly correlated with higher rates of mask use, and that mandating use by all individuals in public spaces is more effective than a less comprehensive mandate for mask use by all public facing employees.

## INTRODUCTION

At the time of writing, the COVID-19 pandemic has killed over 175,000 people in the United States alone^1^. It has drastically impacted economic activity, producing an annualized US GDP contraction of ∼30%, the largest economic contraction since the Great Depression^2^. One tool that can be used to reduce the impact of COVID-19 is wearing a face mask in public. Face masks are a cost-effective tool in contending with respiratory infections^3^. Face mask use reduces the airborne inoculum of COVID-19^4 5 6 7^, and there is some empirical evidence that mask use reduces the spread of COVID-19^8 9 10 11^. The evidence for state policies’ impacts on the use of face masks by the public is more limited^12^. To explore the relationship between state policy and public use of face masks, a previously reported survey of mask use was analyzed and compared between three distinct populations based upon their state-level policies in the United States^13^. This retrospective cohort study was performed to assess the equality of the three populations for reported face mask usage.

## METHODS

A comparative analysis of state level mask policy on public face mask use in the United States was undertaken. Data for mask use was from a previously reported survey^13^. The three groups for comparison were defined by their state level mask policy. The first, states with no state level mask mandate **(None)**. The second, those with state level mandated mask use for public facing employees **(Public)**. The third, those with a state level mandate for all Individuals in public spaces **(All)**. States with a mandate for both public facing Employees and all individuals in public spaces were treated as belonging to the more restrictive All group.

Survey results were calculated for the 3,142 Federal Information Processing Standard (FIPS) counties^14^ in the United States and the District of Columbia from 7/2/2020 until 7/14/2020. The original data on mask usage is from the New York Times and Dynata online survey of 250,000 individuals from the 74,134 US census tracts^13^. The survey data was weighted by age and gender, and survey respondents’ locations were approximated from their ZIP codes to transform raw survey responses into county-level estimates. Then mask-wearing estimates were made for each census tract by taking a weighted average of the 200 nearest responses, with closer responses getting more weight in the average. The county-level estimates were assigned to one of three groups defined by their state mask policy. The comparison is a retrospective cohort study which uses the state level mask mandate as of 7/2/2020, reported in the COVID-19 US state policy database (CUSP)^15^, defining the three groups.

The original survey asked:

> “How often do you wear a mask in public when you expect to be within six feet of another person?”
>
> With responses:
>
> “NEVER”, “RARELY”, “SOMETIMES”, “FREQUENTLY” or “ALWAYS” reported as a percentage of responses for each county.

Additionally, a county Mask Score was calculated as a weighted score for each county.

> Mask Score = (%NEVER*0) + (%RARELY*1) + (%SOMETIMES*2) + (%FREQUENTLY*3) + (%ALWAYS*4)

The three comparison groups were defined by their state policy of face mask use as described above:

> **None (1**,**124 Counties)**: CO, FL, GA, ID, IA, KS, LA, MO, MT, NH, OK, SC, SD, TN, VT and WI
>
> **Public Facing Employees (945 Counties)**: AL, AK, AZ, AR, MN, MS, NE, ND, OH, TX, WV and WY
>
> **All Individuals in Public (1**,**068 Counties)**: CA, CT, DE, DC, HI, IL, IN, KY, ME, MD, MA, MI, NV, NJ, NM, NY, NC, OR, PA, RI, UT, VA and WA

Each group of the three comparison groups were examined for normality with the Shapiro-Francia test for normality, intended for samples of up to 5,000 observations^16^. To visualize the degree of skewness between the three populations a skewness test for normality was determined^17 18^. Analysis of equality-of-populations test was performed using the Kruskal –Wallis test^19 20^ Statistical analysis was done with Stata 16.1^21^.

Potential bias rests principally in the original survey. Respondents were surveyed online and may be representative of subjects more concerned about COVID-19. Additionally, as with any self -report, social desirability impacts reporting. The strength of the study lies in its large sample size and in its geographic reach.

## RESULTS

The Shapiro-Francia test for normal data show that responses in the group **None** was normally distributed (p=0.067) but that the responses in the groups **Public** and **All** were not normally distributed (p ≤ 0.001). The equality of populations test was therefore determined by the Kruskal-Willis test. The populations were statistically significantly different from each other (p ≤ 0.001).

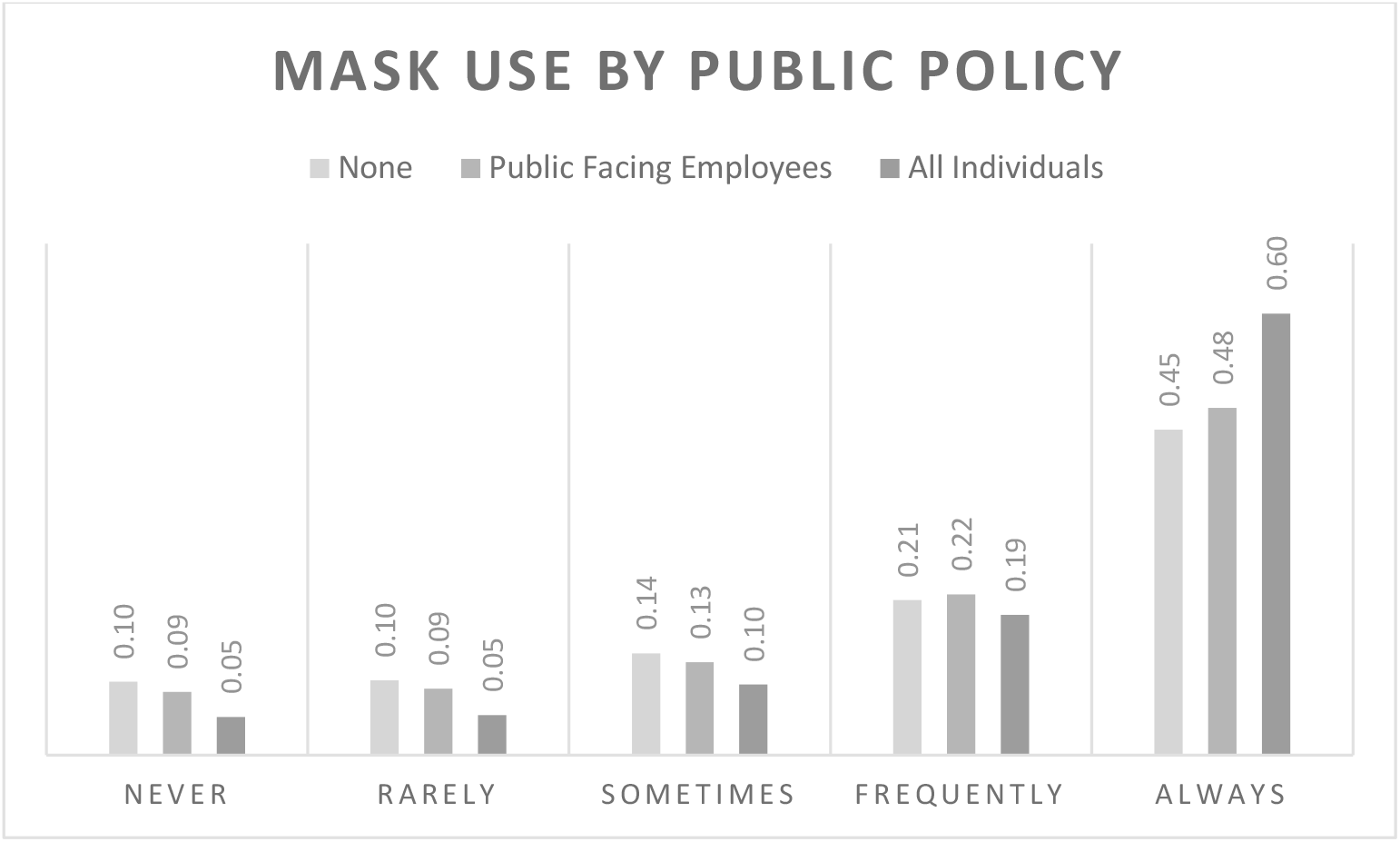

All differences between groups are statistically significant with p ≤ 0.0001

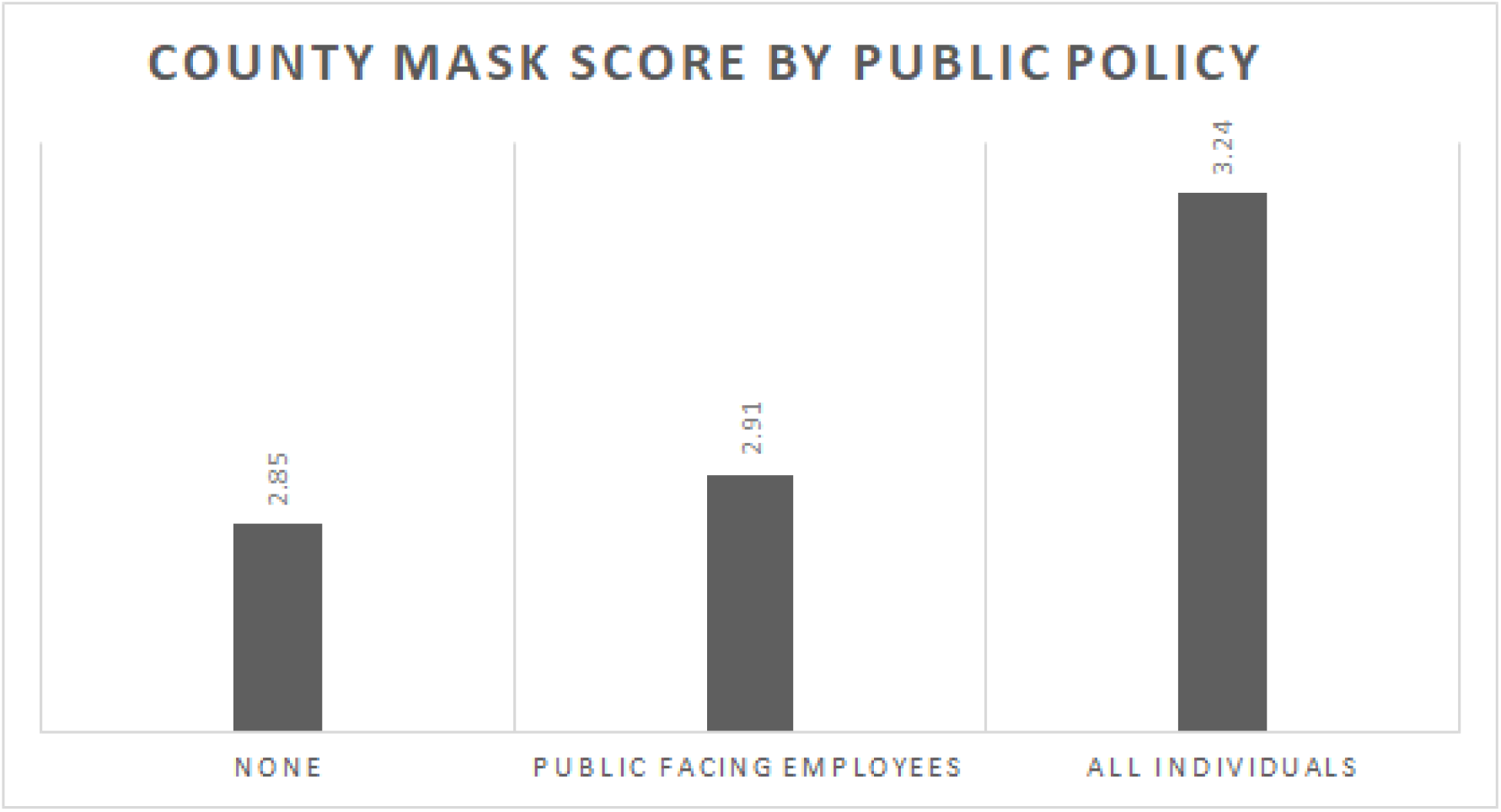

All differences between groups are statistically significant with p ≤ 0.0001. Mask Score for each of the 3124 counties studied was determined by the weighted sum of responses such that MASK SCORE = (%NEVER*0) + (%RARELY*1) + (%SOMETIMES*2) + (%FREQUENTLY*3) + (%ALWAYS*4).

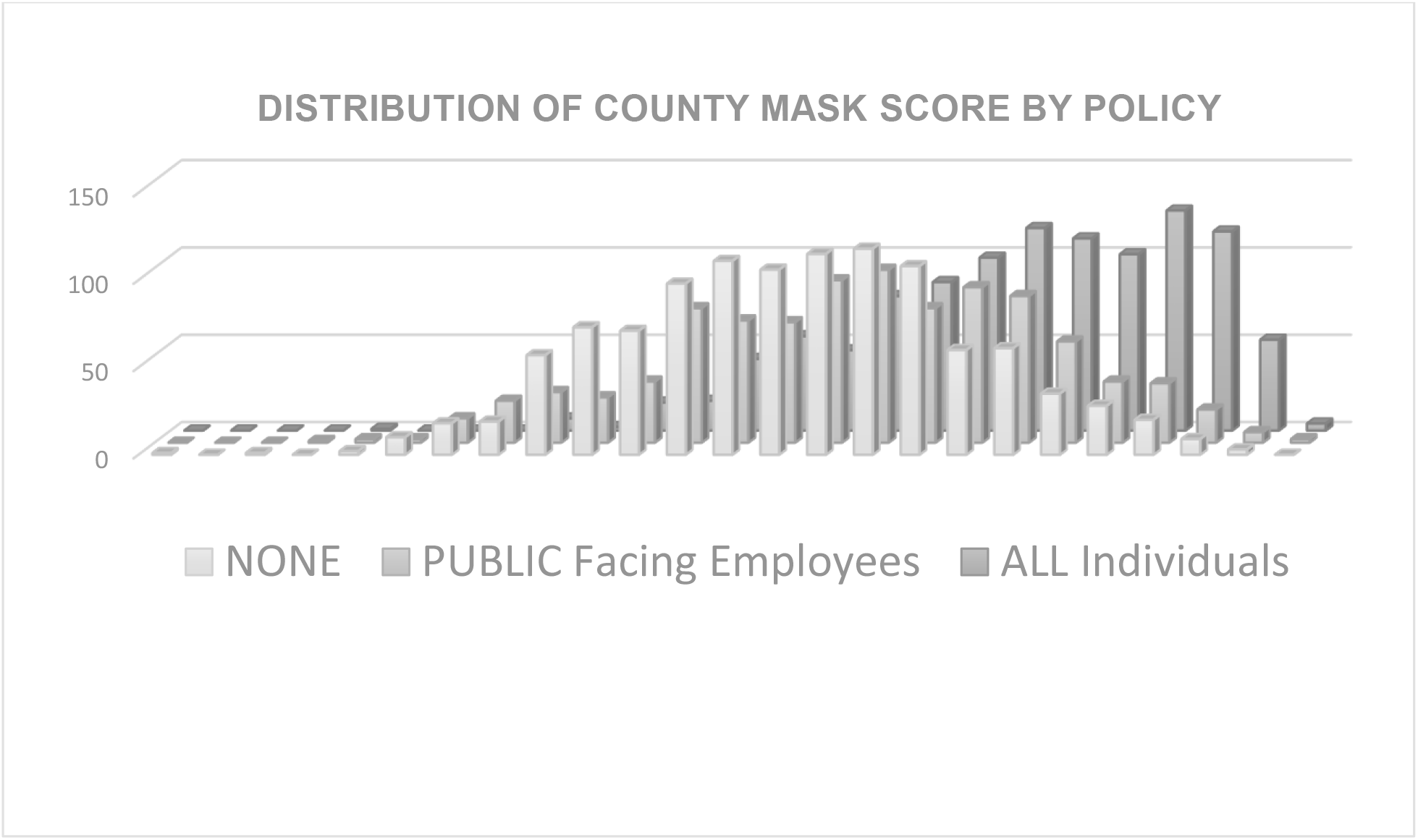

County mask scores show a shift from normal distribution of mask use for people living in counties with no mask mandate (**NONE** skewness of -0.01) to progressively more significant skewness for counties with a mask mandate for all public facing employees (**PUBLIC** skewness of -0.20) and the greatest skewness for the more universal mandate for all individuals in public (**ALL** skewness of -0.57).

## DISCUSSION

Different state policies are related to three different patterns of reported mask use. The more comprehensive the state policy the larger the scale of change – halving the percent of NEVERS and increasing the percent of ALWAYS by a third. The distribution of the responses was normal for the county Mask Score for states with No mandate, but skewed in the states with mask mandates, perhaps indicating an impact of the mandates in shifting attitudes toward face mask use in public.

These findings are consistent with the hypothesis that public policies impact public behaviors relevant to the COVID-19 pandemic. The policy of state level mask mandates appears to increase the use of masks in public. While these results are not sufficient to demonstrate a causal impact on COVID-19 spread, they are suggestive that governmental policies related to mask use may help limit COVID-19 spread. These results further suggest that mask policies may help reduce the health and economic impact of the COVID-19 pandemic. Moreover, these empirical results have clear implications for public policy and support the utility of a national face mask mandate for all individuals in public.

## Data Availability

Re-analysis of previously reported survey in light of state mask policies.

https://github.com/nytimes/covid-19-data/blob/master/mask-use/README.md

https://docs.google.com/spreadsheets/d/1zu9qEWI8PsOI_i8nI_S29HDGHlIp2lfVMsGxpQ5tvAQ/edit#gid=973655443

## AKNOWLEGEMENTS

The author would like to would like to acknowledge Dr. Alan Maloney for his helpful edits and suggestions in reviewing the drafts of this manuscript.

## DISCLOSURES

No relevant disclosures

## Notes

### Competing Interest Statement

The authors have declared no competing interest.

### Clinical Trial

N/A Analysis of public data

### Author Declarations

Publicly available open source data: https://github.com/nytimes/covid-19-data/blob/master/LICENSE https://github.com/USCOVIDpolicy/COVID-19-US-State-Policy-Database

### Summary of Updates

Revised for clarity and grammar. Reformatted per request ID #206326.

